# Coronavirus and birth in Italy: results of a national population-based cohort study

**DOI:** 10.1101/2020.06.11.20128652

**Authors:** Alice Maraschini, Edoardo Corsi, Michele Antonio Salvatore, Serena Donati, ItOSS COVID-19 working group

**Affiliations:** National Centre for Disease Prevention and Health Promotion, Istituto Superiore di Sanità - Italian National Institute of Health, Viale Regina Elena 299, 00161 Rome, Italy; Department of Biomedicine and Prevention, University of Rome Tor Vergata, Viale Montpellier 1, 00133 Rome, Italy; Serena Donati, Alice Maraschini, Ilaria Lega, Paola D’Aloja, Letizia Sampaolo, Michele Antonio Salvatore, Edoardo Corsi, Salvatore Alberico, Paola Casucci, Irene Cetin, Gabriella Dardanoni, Franco Doganiero, Massimo Piergiuseppe Franchi, Livio Leo, Marco Liberati, Mariavittoria Locci Claudio Martini, Federico Mecacci, Alessandra Meloni, Anna Domenica Mignuoli, Luisa Mondo, Enrica Perrone, Luca Ramenghi, Sergio Schettini, Martin Steinkasserer, Saverio Tateo, Vito Trojano

**Keywords:** Pregnancy, Pregnancy complications, Pregnancy outcome, COVID-19, Cohort studies

## Abstract

**Introduction:** The study was implemented to provide guidance to decision-makers and clinicians by describing hospital care offered to women who gave birth with confirmed COVID-19 infection.

**Materials and methods:** National population-based prospective cohort study involving all women with confirmed COVID-19 who gave birth between February 25 and April 22, 2020 in any Italian hospital.

**Results:** The incidence rate of confirmed SARS-CoV-2 infection in women who gave birth was 2.1 per 1000 maternities at a national level and 6.9/1000 in the Lombardy Region. Overall one third of the women developed a pneumonia and 49.7% assumed at least one drug. Caesarean section rate was 32.9%, no mothers nor newborns died. Six percent of the infants tested positive for SARS – CoV-2 at birth.

**Conclusions:** Clinical features and outcomes of COVID-19 in women who gave birth are similar to those described for the general population, most women developing mild to moderate illness.

## Introduction

Following the Chinese epidemic, Italy is currently one of the European countries reporting the highest number of clinical cases. From the H1N1 influenza, SARS, and MERS outbreaks, we learned that pregnant women are at higher risk of developing respiratory complications as well as worse maternal and neonatal outcomes (1). Although there is no conclusive information with regards to an increased susceptibility of pregnant women to the SARS-CoV-2 illness, the currently available data suggest that it is analogous to that of the general population (1,2).

To date, there have been several published case series of deliveries among women affected by COVID-19 (3-6). Available data come primarily from China, and the UK (4).

There is a critical need to develop clinical guidance for obstetric providers and neonatologists, and this should be based on rigorously collected data. The Italian Obstetric Surveillance System (ItOSS) (7) is convenient for establishing a national prospective population-based cohort study on COVID-19 in pregnancy, birth and postpartum in Italy.

This paper describes hospital care of pregnant women with confirmed COVID-19 infection admitted to Italian hospitals for childbirth. Possible transmission of the SARS-CoV-2 virus from mothers to newborns is also addressed.

## Materials and methods

The ItOSS national population-based cohort study collects information on all women who gave birth in any Italian hospital with a SARS-CoV-2 infection confirmed during pregnancy. The COVID-19 diagnosis was detected during pregnancy by reverse transcriptase-polymerase chain reaction (RT-PCR) testing for the SARS-CoV-2 virus through a nasopharyngeal swab and/or by findings from chest X-ray or computed tomography (CT) and/or by antibody response from maternal peripheral blood.

The study’s core outcomes include incidence rate estimate, COVID-19 pneumonia, preterm birth, mode of delivery, invasive respiratory support, intensive care unit (ICU) admission, and maternal and neonatal severe morbidity and mortality.

The ItOSS network of trained reference clinicians working in Italian public and private maternity units covering 91% of total births (7), has been extended to reach nationwide coverage for the present study. Through this system, cases are rapidly notified, and data on maternal and neonatal management collected. Informed consent to the participation is acquired from any woman at study enrolment.

A multidisciplinary expert group of clinicians has revised the data entry form, and its online version has been pre-tested. The form is designed to collect information regarding the woman’s socio-demographic characteristics, medical and obstetrical history, pneumonia diagnosis and treatment, mode of delivery, and maternal and neonatal outcomes.

In case of maternal death from SARS-CoV-2 infection, the ItOSS maternal mortality surveillance system will allow verification and provide further information.

Since this is an observational study, the cohort size depends on the incidence of the disease; therefore, a formal power calculation has not been performed.

The data are collected and processed by personnel responsible for ensuring confidentiality and security. This analysis reports hospitalized cases from February 25 to April 22, 2020, for whom complete data have been received by April 11, 2020.

The incidence rates of women with confirmed SARS-CoV-2 infection who gave birth with a 95% confidence interval (CI) were estimated at a national level, by geographical area, and for the Lombardy Region. Denominator estimates are based on the national Birth Registry providing the most recent available data on deliveries (year 2018) assuming an annual reduction of 0.3% in births. The data analysis focuses on descriptive statistics stratified by the occurrence of pneumonia. Significant differences between the two groups were assessed through the Pearson’s Chi-squared test for categorical variables and Mann-Whitney U test for continuous variables. Data analyses were performed at the Italian National Health Institute using the Statistical Package Stata/MP 14.2.

The Ethics Committee of the Italian National Institute of Health approved the project (Prot. 0010482 CE 01.00, Rome 24/03/2020).

This study has not received any funding.

## Results

From February 25 to April 22, 2020, 146 women who gave birth in any Italian Obstetric Unit with confirmed SARS-CoV-2 infection during pregnancy were notified to ItOSS. The diagnosis of COVID-19 infection was confirmed for 142 patients by RT-PCR testing through nasopharyngeal swab and in 4 cases through chest X-ray.

Out of the total cases, 126 (86.3%) were notified by 5 Regions and 2 Autonomous Provinces located in the North of the country. Among the cases reported in the North, 84 (57.5%) were identified in the Lombardy Region.

Among an estimated 70,343 maternities that took place over the same study period in Italy, the incidence rate of confirmed SARS-CoV-2 infection in women who gave birth was 2.1 per 1000 (CI 95% 1.8-2.4) maternities at a national level, 3.9/1000 (CI 95% 3.2-4.6) in Northern, 1.0/1000 in Central (CI 95% 0.6-1.6), and 0.2/1000 (CI 95% 0.1-0.5) in Southern Italy. The rate in Lombardy was 6.9/1000 (CI 95% 5.5-8.5) maternities.

During the 14 days prior to diagnosis, 41.1% of the women reported having had contact with a probable case (32.9%) or entering health care facilities with confirmed SARS-CoV-2 cases (13.7%).

Tab.1 describes the socio-demographic characteristics of the cohort stratified by COVID-19 pneumonia occurrence. Women’s median age is 32 years (q1-q3=29-36). Women without Italian citizens are 18.5% of the entire cohort, respectively 27.7% and 14.4% of the group with and without pneumonia (p-value=0.049). Previous comorbidities were significantly higher (p-value=0.023) amongst the pneumonia group (34.0%) compared to unaffected women (17.2%), obesity being the most frequent condition. None of the women smoked until the end of pregnancy, and foetal growth restriction was diagnosed in 2.0% of the cases.

**Table 1.** Women’s socio-demographic characteristics by occurrence of COVID-19 pneumonia.

| Characteristics | Total<br>(N=146)<br>n (%) |  | Without COVID-19<br>pneumonia<br>(N=99)<br>n (%) |  | With COVID-19<br>pneumonia<br>(N=47)<br>n (%) |  |
| --- | --- | --- | --- | --- | --- | --- |
| **Maternal age <sup>a</sup>** |  |  |  |  |  |  |
| <30 | 40 | (28.0) | 23 | (24.0) | 17 | (36.2) |
| 30-34 | 53 | (37.1) | 35 | (36.5) | 18 | (38.3) |
| ≥35 | 50 | (35.0) | 38 | (39.6) | 12 | (25.5) |
| **Citizenship** |  |  |  |  |  |  |
| Not Italian | 27 | (18.5) | 14 | (14.1) | 13 | (27.7) |
| Italian | 119 | (81.5) | 85 | (85.9) | 34 | (72.3) |
| **Country of birth** |  |  |  |  |  |  |
| Italy and western Europe | 112 | (76.7) | 83 | (83.8) | 29 | (61.7) |
| East Europe | 10 | (6.8) | 7 | (7.1) | 3 | (6.4) |
| Africa | 9 | (6.2) | 4 | (4.0) | 5 | (10.6) |
| South/Central America | 11 | (7.5) | 4 | (4.0) | 7 | (14.9) |
| Asia | 4 | (2.7) | 1 | (1.0) | 3 | (6.4) |
| **Educational level** |  |  |  |  |  |  |
| ≤8 years | 18 | (12.3) | 10 | (10.1) | 8 | (17.0) |
| >8 years | 86 | (58.9) | 59 | (59.6) | 27 | (57.4) |
| Missing | 42 | (28.8) | 30 | (30.3) | 12 | (25.5) |
| **Previous comorbidities** |  |  |  |  |  |  |
| No | 113 | (77.4) | 82 | (82.8) | 31 | (66.0) |
| Yes | 33 | (22.6) | 17 | (17.2) | 16 | (34.0) |
| *Obesity* | 22 | (15.1) | 11 | (11.1) | 11 | (23.4) |
| *Autoimmune disease* | 4 | (2.7) | 2 | (2.0) | 2 | (4.3) |
| *Diabetes* | 6 | (4.1) | 4 | (4.0) | 2 | (4.3) |
| *Hypertension* | 5 | (3.4) | 1 | (1.0) | 4 | (8.5) |
| *Other* | 3 | (2.1) | 2 | (2.0) | 1 | (2.1) |
| **Smoking in pregnancy** |  |  |  |  |  |  |
| Never | 118 | (80.8) | 78 | (78.8) | 40 | (85.1) |
| Quit before or during pregnancy | 14 | (9.6) | 11 | (11.1) | 3 | (2.1) |
| Missing | 14 | (9.6) | 10 | (10.1) | 4 | (8.5) |

<sup>a</sup> Maternal age has 3 missing values (all in "Without COVID-19 pneumonia" group)

As reported in Tab. 2, multiparas are 69.2% of the cohort and 67.1% of the mothers gave birth vaginally, 25.5% under epidural analgesia. Caesarean section (CS) rate was 32.9% overall, 48.9% among the pneumonia group and 25.3% among the unaffected group (p-value=0.004). COVID-19 indication to CS concerned 7.5% of the entire cohort. General anaesthesia was performed in 5 cases.

**Table 2.** Obstetric characteristics and intra partum care by occurrence of COVID-19 pneumonia.

| Characteristics | Total<br>(N=146)<br>n (%) |  | Without COVID-19<br>pneumonia<br>(N=99)<br>n (%) |  | With COVID-19<br>pneumonia<br>(N=47)<br>n (%) |  |
| --- | --- | --- | --- | --- | --- | --- |
| **Parity** |  |  |  |  |  |  |
| Nulliparae | 45 | (30.8) | 32 | (32.3) | 13 | (27.7) |
| Multiparae | 101 | (69.2) | 67 | (67.7) | 34 | (72.3) |
| **Multiple pregnancy** |  |  |  |  |  |  |
| No | 143 | (97.9) | 97 | (98.0) | 46 | (97.9) |
| Yes | 3 | (2.1) | 2 | (2.0) | 1 | (2.1) |
| **Mode of delivery** |  |  |  |  |  |  |
| Vaginal | 98 | (67.1) | 74 | (74.7) | 24 | (51.1) |
| Elective CS | 12 | (8.2) | 9 | (9.1) | 3 | (6.4) |
| Urgent/emergency CS due to maternal/fetal indication | 25 | (17.1) | 14 | (14.1) | 11 | (23.4) |
| Urgent/emergency CS due to COVID | 11 | (7.5) | 2 | (2.0) | 9 | (19.1) |
| **PPROM<sup>a</sup>** |  |  |  |  |  |  |
| No | 134 | (95.0) | 90 | (94.7) | 44 | (95.7) |
| Yes | 7 | (5.0) | 5 | (5.3) | 2 | (4.3) |
| **Gestational age at birth** |  |  |  |  |  |  |
| ≤32 | 6 | (4.1) | 2 | (2.0) | 4 | (8.5) |
| 33-36 | 22 | (15.1) | 11 | (11.1) | 11 | (23.4) |
| ≥37 | 118 | (80.8) | 86 | (86.9) | 32 | (68.1) |
| Median (q1-q3) | 38 | (37-39) | 38 | (37-40) | 38 | (36-39) |

CS, Cesarean Section; *PPROM*, Preterm Premature Rupture of Membranes; *q1*, 25% percentile; *q3*, 75% percentile
<sup>a</sup> PPRM has 5 missing values (4 in "Without COVID-19 pneumonia" group and 1 in "With COVID-19 pneumonia" group)

Overall, 19.2% of the cohort gave birth preterm, 12.3% due to spontaneous onset of preterm delivery (n=18), and 6.9% due to iatrogenic labour induction (n=3) and urgent/emergency CS (n=7). Preterm birth concerned 31.9% of the women affected by pneumonia compared to 13.1% of the unaffected (p-value=0.007). Among preterm births, the majority were late preterm (Tab.2).

On hospital admission, 28.1% of the women were asymptomatic. The onset of clinical symptoms occurred in 9.5% of the cases on the day of delivery, and in 90.5% before it, the median value being 8 days (range 1-52 days). Overall, fever (47.9%), cough (46.6%), and general weakness (35.6%) were the most common symptoms at presentation. Tab. 3 highlights the higher percentage of symptoms amongst women affected by pneumonia, 31.9% reporting dyspnoea vs. 5.1% of the unaffected (p=0.000).

**Table 3.** Diagnosis and medical therapy by occurrence of COVID-19 pneumonia.

| Characteristics | Total<br>(N=146)<br>n (%) |  | Without COVID-19<br>pneumonia<br>(N=99)<br>n (%) |  | With COVID-19<br>pneumonia<br>(N=47)<br>n (%) |  |
| --- | --- | --- | --- | --- | --- | --- |
| **Symptoms** |  |  |  |  |  |  |
| Fever | 70 | (47.9) | 39 | (39.4) | 31 | (66.0) |
| Cough | 68 | (46.6) | 38 | (38.4) | 30 | (63.8) |
| Tiredness | 52 | (35.6) | 25 | (25.3) | 27 | (57.4) |
| Muscle/joint pain | 28 | (19.2) | 14 | (14.1) | 14 | (29.8) |
| Sore throat | 27 | (18.5) | 16 | (16.2) | 11 | (23.4) |
| Rhinorrhea | 23 | (15.8) | 17 | (17.2) | 6 | (12.8) |
| Dyspnea | 20 | (13.7) | 5 | (5.1) | 15 | (31.9) |
| Headache | 16 | (11.0) | 7 | (7.1) | 9 | (19.1) |
| Vomiting/Diarrhea | 14 | (9.6) | 5 | (5.1) | 9 | (19.1) |
| Chest pain | 5 | (3.4) | 3 | (3.0) | 2 | (4.3) |
| Conjunctivitis | 3 | (2.1) | 3 | (3.0) | 0 | (0.0) |
| No symptoms | 41 | (28.1) | 37 | (37.4) | 4 | (8.5) |
| **Imaging techniques** |  |  |  |  |  |  |
| No exams | 62 | (42.5) | 62 | (62.6) | 0 | (0.0) |
| Chest X-ray | 51 | (34.9) | 26 | (26.3) | 25 | (53.2) |
| Chest CT | 6 | (4.1) | 2 | (2.0) | 4 | (8.5) |
| Lung ultrasound | 6 | (4.1) | 5 | (5.1) | 1 | (2.1) |
| Association of different techniques | 21 | (14.4) | 4 | (4.0) | 17 | (36.2) |
| **Vital signs and laboratory reports** |  |  |  |  |  |  |
| Body temperature >37.5 °C | 34 | (23.3) | 11 | (11.1) | 23 | (48.9) |
| Lymphopenia (<1500 mm3) | 73 | (50.0) | 40 | (40.4) | 33 | (70.2) |
| CRP values >10mg/100ml | 34 | (23.3) | 17 | (17.2) | 17 | (36.2) |
| Oxygen saturation <95% | 17 | (11.6) | 4 | (4.0) | 13 | (27.7) |
| Blood gas test <sup>a</sup> |  |  |  |  |  |  |
| Not performed | 96 | (67.6) | 80 | (82.5) | 16 | (35.6) |
| Performed. normal results | 26 | (18.3) | 13 | (13.4) | 13 | (28.9) |
| Performed. abnormal results | 20 | (14.1) | 4 | (4.1) | 16 | (35.6) |
| **Drugs administered <sup>b</sup>** |  |  |  |  |  |  |
| HCQ + Antivirals + Antibiotics | 20 | (13.8) | 3 | (3.0) | 17 | (37.0) |
| HCQ + Antibiotics | 14 | (9.7) | 6 | (6.1) | 8 | (17.4) |
| Empirical antibiotics | 14 | (9.7) | 12 | (12.1) | 2 | (4.3) |
| HCQ + Antivirals | 8 | (5.5) | 1 | (1.0) | 7 | (15.2) |
| HCQ alone | 8 | (5.5) | 6 | (6.1) | 2 | (4.3) |
| Antivirals + Antibiotics | 5 | (3.4) | 1 | (1.0) | 4 | (8.7) |
| Targeted antibiotics | 2 | (1.4) | 2 | (2.0) | 0 | (0.0) |
| Antivirals alone | 1 | (0.7) | 1 | (1.0) | 0 | (0.0) |
| No pharmacological treatment | 73 | (50.3) | 67 | (67.7) | 6 | (13.0) |
| Antenatal corticosteroids | 9 | (6.2) | 4 | (4.0) | 5 | (10.9) |

CRP, C-reactive protein; CT, Computed Tomography; HCQ, Hydroxychloroquine
<sup>a</sup> Blood gas test has 4 missing values (2 in "Without COVID-19 pneumonia" group and 2 in "With COVID-19 pneumonia" group)
<sup>b</sup> Drugs administered has 1 missing value in "With COVID-19 pneumonia" group

One third of the women developed COVID-19 pneumonia; oxygen saturation <95% and abnormal results of blood gas test concerned respectively 27.7% and 35.6% of the women with pneumonia compared to 4% of the unaffected.

Over 80% of the women who developed pneumonia received at least one pharmacological treatment (Tab.3). Overall, hydroxychloroquine was the most frequently administered drug, alone (5.5%) and in combination with other medical therapies (29.0%). The most frequently adopted therapeutic scheme included hydroxychloroquine in association with antivirals and antibiotics concerning 13.8% of the cohort and 37.0% of the group with pneumonia. Among women with pneumonia, the subgroup at higher risk of worse outcomes – defined by the presence of at least one previous comorbidity or C-reactive protein>10mg/100ml or dyspnoea –received more often the combination of the three drugs. The women belonging to this subgroup, compared to those at lower risk, registered the longest hospital stay (median value 13 days), received more often invasive ventilatory support due to severe morbidity and were admitted more frequently to ICU.

Antenatal corticosteroids for foetal lung maturation were administered respectively to 10.9% and 4.0% of the groups with and without pneumonia.

Severe adverse maternal outcomes have been rare, affecting almost exclusively the group with pneumonia, as described in Tab. 4. Seven women (4.8%) were in critical conditions due to severe morbidity (2 acute respiratory distress syndromes, 1 respiratory failure, 1 preeclampsia, 2 postpartum haemorrhages, and one thrombosis). Invasive ventilatory support concerned 11 women (7.5%), orotracheal intubation 2 mothers (1.4%). ICU admission regarded 7 patients (4.8%), on average for 7 days. None required extracorporeal membrane oxygenation and none died. Two percent of the women were transferred from another hospital, and 18 were still hospitalized at the end of the study period. The median hospital stay was respectively 10 and 4 days for the women with and without pneumonia (p=0.000).

**Table 4.** Maternal outcomes by occurrence of COVID-19 pneumonia.

| Characteristics | Total<br>(N=146) |  | Without COVID-19<br>pneumonia<br>(N=99) |  | With COVID-19<br>pneumonia<br>(N=47) |  |
| --- | --- | --- | --- | --- | --- | --- |
|  | n (%) |  | n (%) |  | n (%) |  |
| Severe morbidity | 7 | (4.8) | 3 | (3.0) | 4 | (8.5) |
| Non invasive respiratory support | 28 | (19.2) | 6 | (6.1) | 22 | (46.8) |
| Invasive respiratory support | 11 | (7.5) | 2 | (2.0) | 9 | (19.1) |
| Orotracheal intubation | 2 | (1.4) | 1 | (1.0) | 1 | (2.1) |
| ICU admission | 7 | (4.8) | 2 | (2.0) | 5 | (10.6) |
| Extracorporeal membrane oxygenation | 0 | (0.0) | 0 | (0.0) | 0 | (0.0) |
| Maternal death | 0 | (0.0) | 0 | (0.0) | 0 | (0.0) |

ICU, Intensive Care Unit

Overall, 2 stillbirths were detected respectively at 30 and 35 weeks of pregnancy (Tab. 5). There were 143 singletons and 3 sets of twins, 85.0% of the newborns weighed ≥2500 gr, 3.4% <1500 gr. Median Apgar index was 9 at 1 minute and 10 at 5 minutes; at 5 minutes, 1 infant had Apgar index <4 (Tab.4). Admission to neonatal intensive care unit (NICU) concerned 23 newborns (15.6%), 18 of whom were preterm, including 6 <32 weeks of gestation. Four infants developed severe morbidity (1 interstitial pneumonia and 3 respiratory distress syndrome). Two were breeches, one affected by spina bifida, and one was macrosoma. None of them tested SARS-CoV-2 positive at birth and, overall, no neonatal death was recorded.

**Table 5.** Fetal and Neonatal outcomes by occurrence of COVID-19 pneumonia.

| Characteristics | Total<br>(N=149) |  | Without COVID-19<br>pneumonia<br>(N=101) |  | With COVID-19<br>pneumonia<br>(N=48) |  |
| --- | --- | --- | --- | --- | --- | --- |
|  | n (%) |  | n (%) |  | n (%) |  |
| Stillbirth | 2 | (1.3) | 1 | (1.0) | 1 | (2.1) |
| Livebirth | 147 | (98.7) | 100 | (99.0) | 47 | (97.9) |
| Neonatal birthweight (g) |  |  |  |  |  |  |
| <1500 | 5 | (3.4) | 3 | (3.0) | 2 | (4.3) |
| 1500-2499 | 17 | (11.6) | 9 | (9.0) | 8 | (17.0) |
| ≥2500 | 125 | (85.0) | 88 | (88.0) | 37 | (78.7) |
| Apgar 1 min > 7 | 128 | (87.1) | 91 | (91.0) | 37 | (78.7) |
| Apgar 5 min > 7 | 140 | (95.2) | 98 | (98.0) | 42 | (89.4) |
| NICU | 23 | (15.6) | 10 | (10.0) | 13 | (27.7) |
| Neonatal morbidity | 4 | (2.7) | 1 | (1.0) | 3 | (6.4) |
| Neonatal death | 0 | (0.0) | 0 | (0.0) | 0 | (0.0) |
| Neonatal positive SARS-CoV-2 test |  |  |  |  |  |  |
| No | 138 | (93.9) | 94 | (94.0) | 44 | (93.6) |
| Positive test <24 hrs of age | 5 | (3.4) | 4 | (4.0) | 1 | (2.1) |
| Positive test ≥ 24 hrs of age | 4 | (2.7) | 2 | (2.0) | 2 | (4.3) |

NICU, Neonatal Intensive Care Unit

Nine newborns (6.1%) tested positive for SARS – CoV-2, 5 were tested on the day of delivery, 1 the day after, and 3 after 6-9 days from birth. Out of the 5 newborns with positive swabs collected within 24 hours from birth, 4 were delivered vaginally and 1 by pre-labour CS. Three of the positive infants were admitted to NICU, and none of them developed a severe illness.

## Discussion

Women’s mean age is 32.5 years (SD=5.5) in line with the mean age of women giving birth in Northern Italy (8,9). Thirty-two percent of the cohort developed COVID-19 pneumonia; the previous comorbidity being significantly associated (p-value=0.023). 0ver 80% of the women who developed pneumonia received at least one pharmacological treatment, hydroxychloroquine being the most largely prescribed. Preterm births concerned 19.2% of the cohort with statistically different figures (p-value = 0.007) between the pneumonia group (31.9%) and the other (13.1%). CS rate was 32.9%, 7.5% due to COVID-19 indication. Respectively 4.8% of the mothers and 2.7% of the newborns developed severe morbidity. No mothers nor newborns died. Six percent of the infants tested positive for SARS – CoV-2 at birth.

The incidence rates of SARS-CoV-2 positive women who gave birth in Italy (2.1/1000), in the North (3.9/1000), the Centre (1/1000) and the South of the country (0.2/1000) as well as in the Lombardy Region (6.9/1000) reflect the same variation in circulation of the virus among geographical areas as that detected in the general population (10). This observation impacts on the present different seroprevalence and susceptibility of the population. In the current phase, following the lockdown, the early identification of new cases and their contact traceability and isolation will be decisive for the containment of the virus circulation.

Compared to the reference population of women giving birth in Northern Italy (8,9), the higher proportion of multiparas (69% vs 50%) confirms the hypothesis of greater circulation of the virus in families with children who are often asymptomatic.

Clinical features of the detected pneumonia are similar to those described by previous studies (2,6,11,12). The majority of cases are mild/moderate, and similarly to the UKOSS study (4) previous comorbidities are significantly associated with pneumonia (p-value= 0.023).

In Italy, black and minority ethnicities are present in a lower proportion than in the UK where a significative association with COVID-19 has been detected (4). Hospitalized women without Italian citizenship develop significantly more often pneumonia than the Italians (p-value=0.049), probably due to delayed access to healthcare services.

Preterm birth, which is one of the feared outcomes of COVID-19, was overall 21.2% in the UK cohort (4), 17.4% in the WHO review (6), and 19.2% in Italy compared to the 7% North Italian figure (8,9).

Mothers requiring critical care were 9% in the UK (4) and 7.5% in Italy. NICU admissions concerned respectively 26% in the UK (4), 6.2% of the infants described by the WHO review (6), and 4.8% of the Italian newborns (Tab.4) but this variability could be related to different local admission policies to NICU regarding quarantine or neonatal observation. The UK cohort reported 5 maternal and 2 neonatal deaths compared to zero deaths reported in Italy and China. The interpretation of these differences is not straightforward even though the lower prevalence of minority ethnicities could play a role as well as the different pattern of drug prescriptions.

The data are preliminary and collection is still ongoing; nevertheless, they confirm a better course of the disease compared to H1N1flu and SARS and MERS epidemics (1,2).

The observation that almost 60% of the enrolled women did not have risky contacts during the 14 days prior to symptom onset raises the challenge of the impact of asymptomatic infections and the opportunity to consider screening policies for pregnant women at hospital admission, currently available in few Italian Regions and/or hospitals.

The proportion of women with pneumonia receiving pharmacological treatment in this study is high, 73.9% receiving hydroxychloroquine alone or in association with antivirals and/or empirical antibiotics (Tab.3). Lopinavir amongst antivirals is the most frequently used, probably due to the experience of its use in HIV positive women. Conversely, among the UKOSS cohort hydroxychloroquine was not administered at all and 2% of the women received antivirals (4). The ItOSS data do not confirm that this medication has resulted in a benefit compared to standard care. Before a treatment protocol can be confidently used in clinical settings, evidence on its safety and effectiveness is needed. Antenatal corticosteroids for fetal lung maturation have been prescribed as recommended (2) to 4 women <34 weeks and to 5 women at 34-35 weeks of gestation. The detected rate of CS in the Italian cohort is 32.9%, higher compared to the rate of the Northern Regions (26%) but considerably lower compared to 59% of the UK cohort (4), 73.5% reported by the WHO review (6), and 85% of the Chinese series (3-6). As reported in the UK, the majority of CS indications were not due to SARS-CoV-2 infection, which is not in itself necessarily an indication for delivery, nor for CS. The proportion of CS due to COVID-19 concerned in fact a minority of the total surgeries, 7.5% of Italian and 16% of UK cohort. Although international agencies are unequivocal in claiming that the disease is not an indication for CS and that the protection of birth physiology is a priority (2,13,14), clinical practice seem not to follow the current recommendations. Italy, which has historically recorded higher CS rates compared to the UK, on this occasion didn’t show the same significant increase in CS as in other countries despite the early onset of the epidemic. An issue still highly debated concerns the possibility of mother to foetus transmission of SARS-CoV-2 virus. The WHO review shows that 6.6% of the newborns tested as suspected to have COVID-19, the UK cohort reports 5% of positive infants, and the ItOSS cohort 6%. Although evidence is sparse, vertical transmission cannot be excluded (15,16) but it appears to be rare and, for the most part, babies, who must be carefully monitored, have a good prognosis.

Many case reports and small case series have already been published on COVID-19 in pregnancy; however, there is a lack of population-based data that could allow incidence rates to be estimated and unbiased characteristics and outcomes to be described and compared (4). The scientific community, as well as international journal peer review procedures, should better support the development and the dissemination of studies adopting population-based approaches to properly inform clinicians and decision-makers.

Pregnant women are often not included in clinical trials on drugs (17). However, the good news is that recently the EMA and Health Canada under the aegis of the International Coalition of Medicines Regulatory Authorities have agreed on three priority areas for cooperation on observational research during the outbreak of COVID-19 (18). One area is devoted to research in pregnancy in order to examine the impact of both Coronavirus disease and the use of drugs on pregnant women infected with SARS-CoV-2 and their unborn babies. The different practices in prescribing hydroxychloroquine and antivirals observed in the UK and Italy promotes a reflection on the determining factors that guide clinicians in deciding whether and to what extent they should confidently prescribe drugs for which conclusive evidence is still unavailable.

The study’s strengths are the national population-based prospective design and the opportunity to analyse data from the beginning of the epidemic. Another asset is the wealth of information contained in the data collection form. Limitations include the analysis of preliminary data while the pandemic is still underway and the constraints linked to the impossibility of generalizing the results without taking into account the different prevalence of the condition by geographical area. The lack of information regarding women infected in the early stages of pregnancy is also a limit of the study, but the ItOSS will follow-up affected women currently in the first trimester of pregnancy.

## Conclusions

The clinical presentation of COVID-19 in women who gave birth appears to be similar to the general population. The lesson learned by reviewing hospital care offered to affected Italian women who gave birth, confirmed the current priority of physical distancing measures, the urgent need for stronger evidence on the safety and effectiveness of medical therapy and a continued commitment to face the challenge of respecting and protecting childbirth physiology. These findings confirm the primary importance of comparisons among population-based cohorts to support health professionals and decision-makers with evidence-based recommendations.

## Data Availability

All data referred to in the manuscript are available in the Istituto Superiore di Sanita

## Acknowledgements

We thank Silvia Andreozzi and Mauro Bucciarelli for their valuable technical support and assistance to the operation of the web-based data collection system. We thank Clarissa Bostford for language editing.

Our heartfelt thanks go to all the clinicians working in the national network of maternity units (Appendix) for the assistance offered to women and for collecting the data, we thank all women who agreed to participate in the study.

## Individual contribution to the manuscript

Alice Maraschini: Conceptualization, Methodology, Software, Formal Analysis, Writing-Review & Editing Edoardo Corsi: Methodology, Investigation, Data Curation, Writing-Review & Editing, Project Administration, Antonio Michele Salvatore: Methodology, Software, Formal Analysis, Writing-Review & Editing Serena Donati: Conceptualization, Methodology, Formal Analysis, Writing Original Draft, Supervision

Ilaria Lega:Investigation, Writing-Review & Editing

Paola D’Aloja: Investigation, Writing-Review & Editing

Letizia Sampaolo: Resourches, Writing-Review & Editing

Paola Casucci: Investigation, Data Curation

Irene Cetin: Investigation, Data Curation,

Gabriella Dardanoni: Investigation, Data Curation

Franco Doganiero: Investigation

Massimo Piergiuseppe Franchi: Investigation,

Data Curation Livio Leo: Investigation, Data Curation

Marco Liberati: Investigation, Data Curation

Mariavittoria Locci: Investigation, Data Curation

Claudio Martini: Investigation, Data Curation

Federico Mecacci: Investigation, Data Curation

Alessandra Meloni: Investigation, Data Curation

Anna Domenica Mignuoli: Investigation, Data Curation

Luisa Mondo:Investigation, Data Curation

Enrica Perrone: Investigation, Data Curation,

Writing Review & Editing,

Luca Ramenghi: Investigation, Data Curation

Sergio Schettini: Investigation, Data Curation

Martin Steinkasserer: Investigation, Data Curation,

Saverio Tateo:Investigation, Data Curation

Vito Trojano: Investigation, Data Curation.

## Source of financial support

This study has not received any financial support.

**The ItOSS COVID-19 working group**

Serena Donati^1^, Alice Maraschini^1^, Ilaria Lega^1^, Paola D’Aloja^1^, Letizia Sampaolo^1^, Michele Antonio Salvatore^1^, Edoardo Corsi^2^, Salvatore Alberico^3^, Paola Casucci^4^, Irene Cetin^5^, Gabriella Dardanoni^6^, Franco Doganiero^7^, Massimo Piergiuseppe Franchi^8^, Livio Leo^9^, Marco Liberati^10^, Mariavittoria Locci^11^, Claudio Martini^12^, Federico Mecacci^13^, Alessandra Meloni^14^, Anna Domenica Mignuoli^15^, Luisa Mondo^16^, Enrica Perrone^17^, Luca Ramenghi^18^, Sergio Schettini^19^, Martin Steinkasserer^20^, Saverio Tateo^21^, Vito Trojano^22^

^1^Centre for Disease Prevention and Health Promotion, Istituto Superiore di Sanità - Italian National Institute of Health, Rome, Italy

^2^Department of Biomedicine and Prevention, University of Rome Tor Vergata, Rome, Italy

^3^IRCCS Burlo Garofolo, Trieste, Italy ^4^Umbria Region, Perugia, Italy ^5^University of Milan, Milan, Italy ^6^Sicily Region, Palermo, Italy

^7^ Civil Hospital Antonio Cardarelli, Campobasso, Italy

^8^University of Verona, Verona, Italy

^9^ Beauregard Hospital Valle D’Aosta, Aosta, Italy

^10^University Gabriele d’Annunzio, Chieti-Pescara, Italy

^11^University of Naples Federico II, Naples, Italy

^12^Marche Region, Ancona, Italy

^13^University of Florence, Florence, Italy

^14^Sardegna Region, Cagliari, Italy

^15^Calabria Region, Reggio Calabria, Italy

^16^ASL TO3, Turin, Italy

^17^Emilia-Romagna Region, Bologna, Italy

^18^IRCSS Giannina Gaslini, Genoa, Italy

^19^ S. Carlo Hospital, Potenza,Italy

^20^ Central Hospital of Bozen, Bozen, Italy

^21^Santa Chiara Hospital, Trento, Italy

^22^Mater Dei Hospital, Bari, Italy

## References

1. Schwartz DA, Graham AL. Potential Maternal and Infant Outcomes from (Wuhan) Coronavirus 2019-nCoV Infecting Pregnant Women: Lessons from SARS, MERS, and Other Human Coronavirus Infections. Viruses. 2020;12(2):194. Published 2020 Feb 10. doi:10.3390/v12020194

2. Royal College of Obstetricians and Gynaecologists, Royal College of Midwives, Royal College of Paediatrics and Child Health, Public Health England and Health Protection Scotland. Coronavirus (COVID-19) Infection in Pregnancy. Information for healthcare professionals. Version 9: Published Wednesday 13 May 2020 [cited2020 May 28]. Available from: <https://www.rcog.org.uk/globalassets/documents/guidelines/2020-05-13-coronavirus-covid-19-<infection-in-pregnancy.pdf

3. Della Gatta AN, Rizzo P, Pilu G, Simonazzi G. COVID-19 during pregnancy: a systematic review of reported cases [published online ahead of print, 2020 Apr 18]. Am J Obstet Gynecol. 2020;S0002-9378(20)30438-5. doi:https://doi.org/10.1016/j.ajog.2020.04.013

4. Knight M, Bunch K, Vousden N, Morris E, Simpson N, Gale C, O’Brien P, Quigley M, Blocklehurst P, Kurinczuk JJ. Characteristics and outcomes of pregnant women admitted to hospital with confirmed SARS-CoV-2 infection in the UK: national population based cohort study. BMJ. 2020;369:m2107. doi: https://doi.org/10.1136/bmj.m2107

5. Yan J, Guo J, Fan C, et al. Coronavirus disease 2019 (COVID-19) in pregnant women: A report based on 116 cases [published online ahead of print, 2020 Apr 23]. Am J Obstet Gynecol. 2020;S0002-9378(20)30462-2. doi:10.1016/j.ajog.2020.04.014

6. World Health Organization Collaborating Centre for Women’s Health University of Birmingham. COVID-19 in pregnancy (PregCOV-19 LRS) [cited2020 May 28]. Available from: https://www.birmingham.ac.uk/research/who-collaborating-centre/pregcov/index.aspx

7. EpiCentro [Internet]. 2020 Feb 20 [cited2020 May 28]. ItOSS – Italian Obstetric Surveillance System. Italian. Available from: <https://www.epicentro.iss.it/itoss/>

8. Campi R, Cartabia M, Miglio D, Bonati M. Certificato di Assistenza al Parto (CedAP) Regione Lombardia. Analisi dell’evento nascita anno 2017 - Laboratorio per la Salute Materno Infantile, Dipartimento di Salute Pubblica, IRCCS-Istituto di Ricerche Farmacologiche Mario Negri Milano: Lombardia Region 2019 [cited2020 May 28]. Italian. Available from: <https://www.dati.lombardia.it/dataset/Rapporto-Cedap/v74r-mqr5>

9. Perrone E, Formisano D, Gargano G et al. [Birth in Emilia-Romagna. 2018] Bologna: Emilia-Romagna Region, 2019 [cited2020 May 28]. Italian. Available from: http://www.saperidoc.it/flex/cm/pages/ServeBLOB.php/L/IT/IDPagina/349

10. The COVID-19 Task force of the Department of Infectious Diseases and the IT Service Istituto Superiore di Sanità - Epidemics COVID-19, National update 28th April 2020 [cited2020 May 28]. Italian. Available from: <https://www.epicentro.iss.it/coronavirus/bollettino/Bollettino-sorveglianza->integrata-COVID-19_28-aprile-2020.pdf

11. Yang Z, Wang M, Zhu Z, Liu Y. Coronavirus disease 2019 (COVID-19) and pregnancy: a systematic review [published online ahead of print, 2020 Apr 30]. J Matern Fetal Neonatal Med. 2020;-4. doi:10.1080/14767058.2020.1759541

12. Arabi S, Vaseghi G, Heidari Z, et al. Clinical characteristics of COVID-19 infection in pregnant women: a systematic review and meta-analysis. medRxiv 2020.04.05.20053983 [preprint]. 2020 [cited2020 May 28]. Available from: https://doi.org/10.1101/2020.04.05.20053983.

13. Poon LC, Yang H, Dumont S, et al. ISUOG Interim Guidance on coronavirus disease 2019 (COVID-19) during pregnancy and puerperium: information for healthcare professionals - an update [published online ahead of print, 2020 May 1]. Ultrasound Obstet Gynecol. 2020;10.1002/uog.22061. doi:10.1002/uog.22061

14. World Health Organization. Clinical management of severe acute respiratory infection when novel coronavirus (nCoV) infection is suspected – Interim guidance – 13 March 2020 [cited2020 May 28]. Available from: <https://www.who.int/docs/default-source/coronaviruse/clinical-management-of-novel-cov.pdf>

15. Dong L, Tian J, He S, et al. Possible Vertical Transmission of SARS-CoV-2 from and Infected Mother to Her Newborn [published online ahead of print, 2020 Mar 26]. JAMA. 2020;323(18):1846–1848. doi:10.1001/jama.2020.4621

16. Patanè L, Morotti D, Giunta MR, et al. Vertical transmission of COVID-19: SARS-CoV-2 on the fetal side of the placenta in pregnancies with COVID-19 positive mothers and neonates at birth. Am J Obstet Gynecol MFM. 2020;100145. doi:10.1016/j.ajogmf.2020.100145

17. Costantine MM, Landon MB, Saade GR. Protection by exclusion: another missed opportunity to include pregnant women in research during the coronavirus disease 2019 (COVID-19) pandemic [published online ahead of print, 2020 Apr 24]. Obstet Gynecol. 2020. doi:10.1097/AOG.0000000000003924

18. European Medicines Agency [Internet]. EMA/275870/2020 [cited2020 May 28]. Global regulators commit to cooperate on observational research in the context of COVID-19. Available from: https://www.ema.europa.eu/en/news/global-regulators-commit-cooperate-observational-research-context-covid-19

